# Optimizing Parkinson’s Disease progression scales using computational methods

**DOI:** 10.1101/2025.07.31.25332494

**Authors:** Assaf Benesh, Roy N. Alcalay, Anat Mirelman, Ron Shamir

## Abstract

Parkinson’s disease is a highly heterogeneous condition with symptoms spanning motor and non-motor domains. Clinical scales like the Movement Disorder Society’s Unified Parkinson’s Disease Rating Scale (MDS-UPDRS), are standard in clinical trials where disease progression is monitored. They rely on summing item values, assuming uniform item importance and score increments.

Here we propose a novel data-driven approach to optimize weights for such scales — so that total scores better reflect the underlying disease severity. By leveraging large-scale longitudinal data from the Parkinson’s Progression Markers Initiative (PPMI), our methods identified which items (and value increments) most strongly indicate PD progression, down-weighting or excluding less informative items. The learned weights substantially improve the monotonic relationship between total scores and clinical progression. We validated our weights using both held-out PPMI data and an independent dataset (BeaT-PD), demonstrating their robustness. Applying such weights in clinical trials may increase power and reduce the required sample size [**1**].

## 1 Introduction

Parkinson’s disease (PD) is a complex, progressive neurological disorder characterized by a range of motor and non-motor symptoms. The most commonly used assessment in PD is the Movement Disorder Society’s Unified Parkinson’s Disease Rating Scale (MDSUPDRS) [**2**], a 65-item scale divided into four parts. Each item has five possible answers numbered 0 to 4, reflecting increasing severity. Although thoroughly validated [**3**–**5]** and widely accepted [**6**], MDS-UPDRS exhibits several limitations. First, the total score is obtained by summing item scores, assuming that all items—and all increments within items—are equally informative. For instance, a score of 2 on two different items could have markedly different clinical implications, yet both add the same amount to the total score. Similarly, increasing an item’s score from 0 to 2 has the same effect on the total score as increasing it from 2 to 4, although these increments have different clinical significance. Second, medication-induced fluctuations in the score may not reflect disease progression and obscure the underlying trajectory of PD. Lastly, administering the full scale is time-consuming and some items may not consistently contribute to tracking disease progression.

Due to these reasons, a more robust measure of disease trajectory is needed — one that captures underlying progression better and is also less affected by medication regimens. Moreover, identifying and discarding redundant or minimally informative questions can streamline patient evaluations, reducing both clinical burden and patient fatigue. This work aimed to optimize the weighting of MDS-UPDRS (and related) scale items in order to produce a more accurate and concise PD progression index, capture biology better, and help reduce recruitment needs for clinical trials.

We formulated an optimization problem that seeks weights yielding a score that increases as patients progress, thereby providing a more accurate representation of the true disease state, which we assume progresses monotonically (but not necessarily linearly [**7**]) over time [**8**–**10]**. Concretely, for each MDS-UPDRS question we allowed assigning different weights to the increments between answers (0 to 1, 1 to 2, 2 to 3, 3 to 4), and possibly different weights for different questions, and leveraged computational methods (e.g., linear or integer programming) to choose the weights such that the longitudinal monotonicity of the data is maximized. This data-driven approach enabled us to (1) discover the relative importance of different items and scores, (2) reduce the influence of medication-induced fluctuations by focusing on genuine signs of progression, and (3) minimize redundancy by identifying and down-weighting less informative items.

We constructed six indexes corresponding to different exact or approximate formulations of the optimization problem, named MeanDiff, MeanDiff-W, MeanDiff-QP, MeanDiff-SV, Cons and Cons-Int. The MeanDiff variants aim to maximize the overall score increase between later and earlier examinations from the same patient, while the two Cons versions aim to optimize *consistency*, defined as the proportion of exam pairs from the same patient in which the later exam attains a higher score. We used data from the Parkinson’s Progression Markers Initiative (PPMI) [**11**] to develop the indexes and validated them on held-out PPMI data, external progression criteria and an external cohort of PD patients obtained from the BeaT-PD project (204-16TLV) [**12**]. The resulting indexes offer greater accuracy and more concise measurement instruments.

## 2 Results

### 2.1 Comparing the performance of the different approaches

Table 1 presents the characteristics of the final PPMI cohort after data cleaning.

**Table 1.**
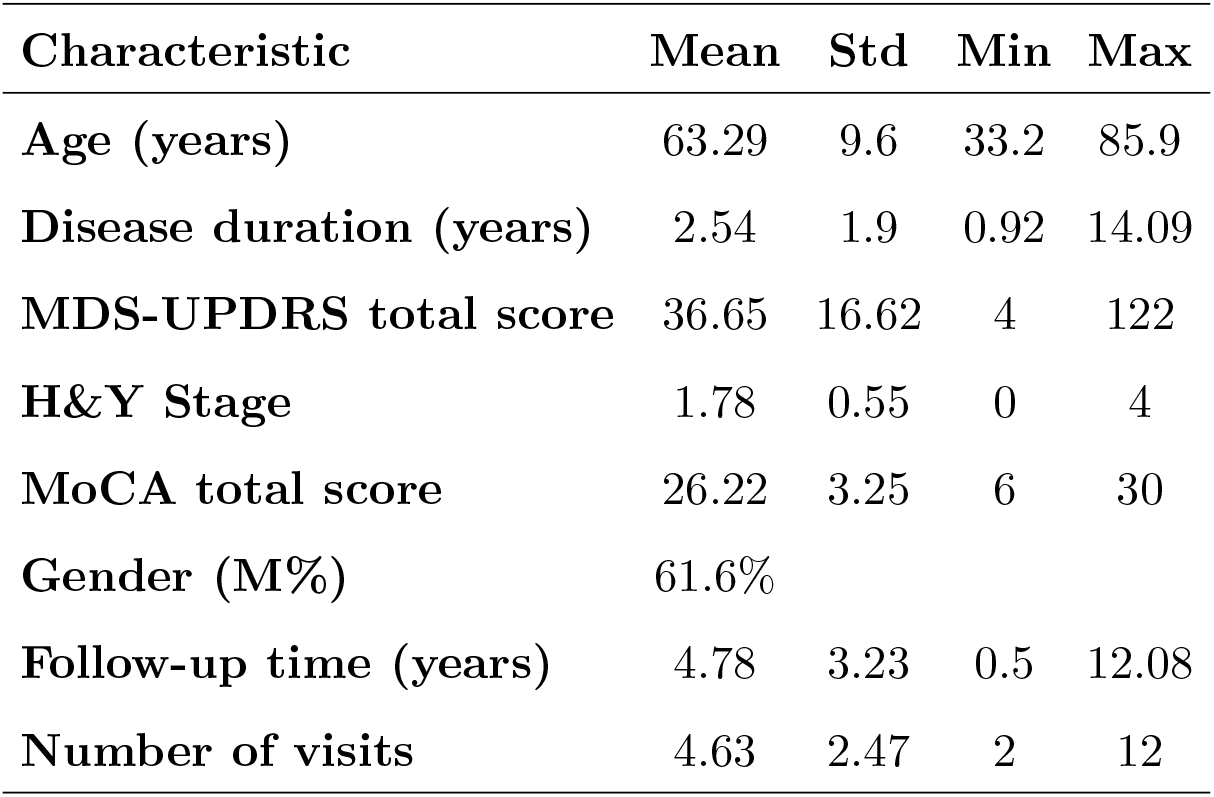
PPMI participants characteristics. Results are shown for PPMI cohort after filtering (3,295 examinations for 711 PD patients). The first five characteristics listed correspond to each patient’s first visit included in the analysis. MDS-UPDRS: Movement Disorder Society’s Unified Parkinson’s Disease Rating Scale. MoCA: Montreal Cognitive Assessment. H&Y: Hoehn and Yahr.

| Characteristic | Mean | Std | Min | Max |
| --- | --- | --- | --- | --- |
| Age (years) | 63.29 | 9.6 | 33.2 | 85.9 |
| Disease duration (years) | 2.54 | 1.9 | 0.92 | 14.09 |
| MDS-UPDRS total score | 36.65 | 16.62 | 4 | 122 |
| H&Y Stage | 1.78 | 0.55 | 0 | 4 |
| MoCA total score | 26.22 | 3.25 | 6 | 30 |
| Gender (M%) | 61.6% |  |  |  |
| Follow-up time (years) | 4.78 | 3.23 | 0.5 | 12.08 |
| Number of visits | 4.63 | 2.47 | 2 | 12 |

Table 2 shows the results when all items were used, as well as the results from applying the original weights of the complete MDS-UPDRS, its individual sections, and MoCA. The new methods outperformed the MDS-UPDRS and its parts as well as MoCA, with MeanDiff-QP performing best.

**Table 2.**
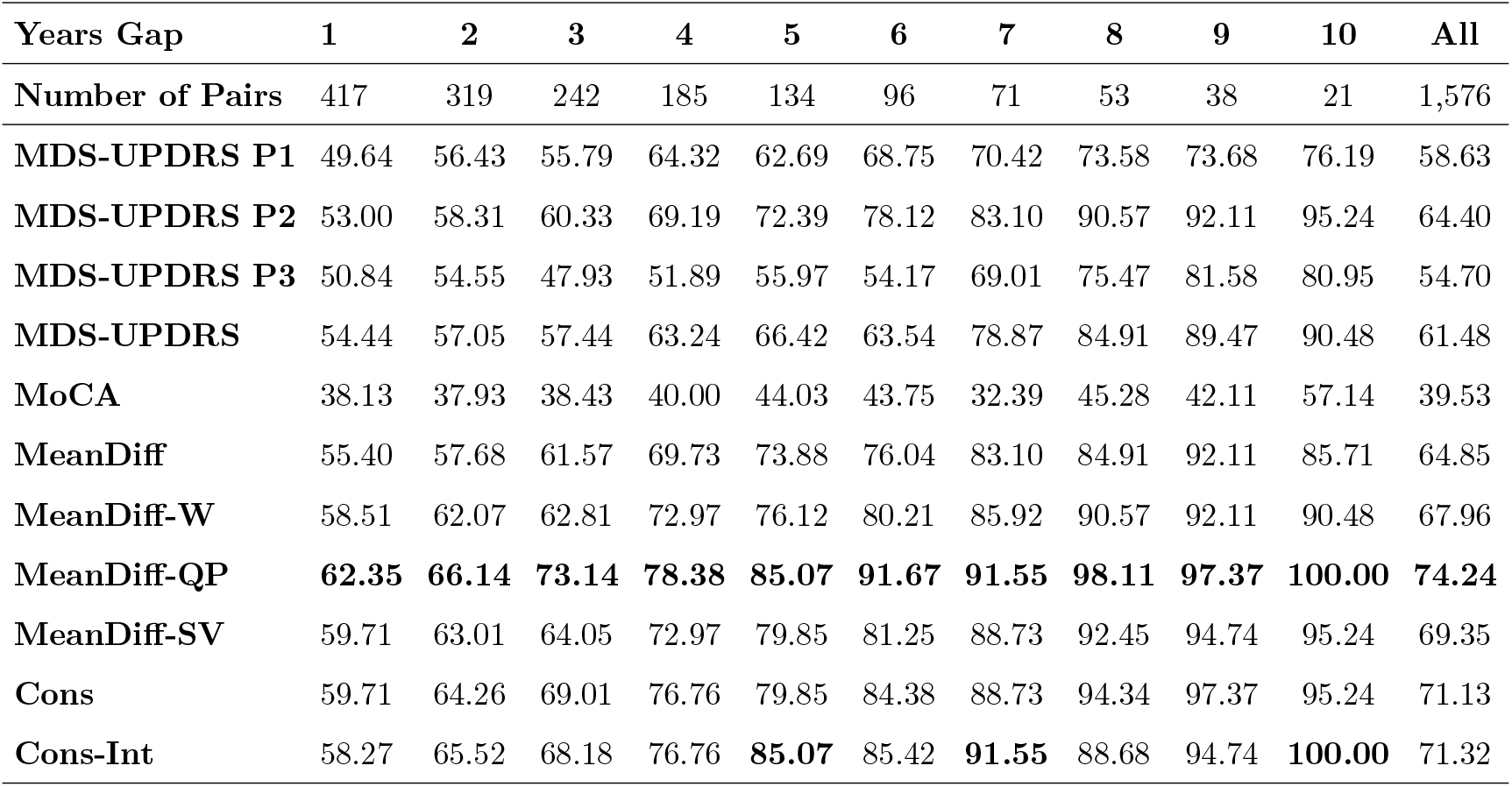
Performance comparison when all MDS-UPDRS and MoCA items are used. The table shows the percentage of consistent pairs of visits for each method, for different time gaps between the visits. Time gaps are rounded to the closest year. The number in bold shows the best performer for each gap. The last column gives the weighted average percentage of consistent pairs. MDS-UPDRS: Movement Disorder Society’s Unified Parkinson’s Disease Rating Scale. MoCA: Montreal Cognitive Assessment.

When limited to self-reported items (Table 3), a similar advantage of the new scale is observed. MeanDiff-QP performs on par with the Cons method, with the latter being slightly better for shorter time gaps.

**Table 3.** Performance comparison when only the self-reported items in MDS-UPDRS are used. See the caption of Table 2 for details. MoCA and MDS-UPDRS part 3 are excluded as they do not contain self-reported items.

| Years Gap | 1 | 2 | 3 | 4 | 5 | 6 | 7 | 8 | 9 | 10 | All |
| --- | --- | --- | --- | --- | --- | --- | --- | --- | --- | --- | --- |
| Number of Pairs | 417 | 319 | 242 | 185 | 134 | 96 | 71 | 53 | 38 | 21 | 1,576 |
| MDS-UPDRS P1 | 46.76 | 52.66 | 58.68 | 61.62 | 57.46 | 65.62 | 71.83 | 75.47 | 76.32 | 66.67 | 56.66 |
| MDS-UPDRS P2 | 53.00 | 58.31 | 60.33 | 69.19 | 72.39 | 78.12 | 83.10 | 90.57 | 92.11 | 95.24 | 64.40 |
| MDS-UPDRS | 53.96 | 61.13 | 66.94 | 74.05 | 74.63 | 72.92 | 85.92 | 90.57 | 94.74 | **100.00** | 66.94 |
| MeanDiff | 54.44 | 61.76 | 66.94 | 72.43 | 76.12 | 73.96 | 84.51 | 92.45 | 94.74 | **100.00** | 67.20 |
| MeanDiff-W | 54.20 | 61.13 | 66.12 | 72.43 | 77.61 | 76.04 | 84.51 | 90.57 | 94.74 | **100.00** | 67.07 |
| MeanDiff-QP | 58.99 | 61.13 | **71.90** | **76.22** | 82.09 | 84.38 | **91.55** | **96.23** | **97.37** | **100.00** | 71.13 |
| MeanDiff-SV | 58.51 | 64.89 | 67.36 | 74.05 | 76.12 | 80.21 | 88.73 | 94.34 | 94.74 | **100.00** | 69.48 |
| Cons | **60.91** | **67.71** | 68.60 | **76.22** | **83.58** | 87.50 | **91.55** | 94.34 | 92.11 | 90.48 | **72.46** |
| Cons-Int | 54.68 | 64.58 | 67.77 | 74.59 | 80.60 | **88.54** | 90.14 | 92.45 | 92.11 | 100.00 | 69.67 |

Figure 1 compares the performance of MeanDiff-QP and MDS-UPDRS when all items are used and when only self-reported items are used. Remarkably, in both cases our optimized method shows better consistency compared to MDS-UPDRS across all time gaps. Moreover, the self-reported version is almost as good as what we can get with all items.

**Figure 1.**
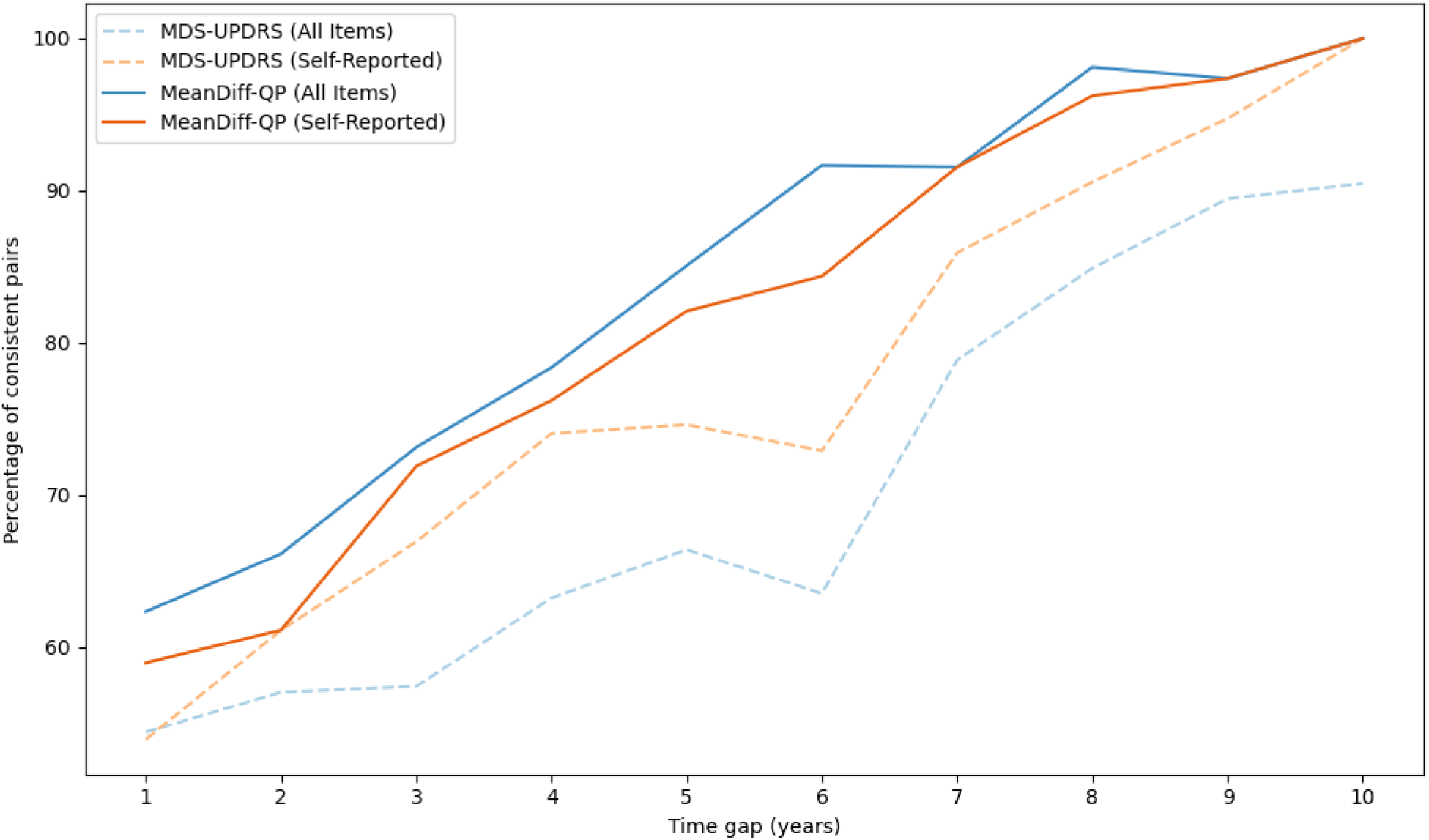
Percentage of consistent pairs for MDS-UPDRS and our MeanDiff-QP scale in various time gaps. MDS-UPDRS: Movement Disorder Society’s Unified Parkinson’s Disease Rating Scale.

### 2.2 Reducing the number of items

Figure 2 shows, for each method, its consistency and the number of non-zero items used. In Figure 2A all items were considered, and in Figure 2B only the self-reported items were allowed.

**Figure 2.**
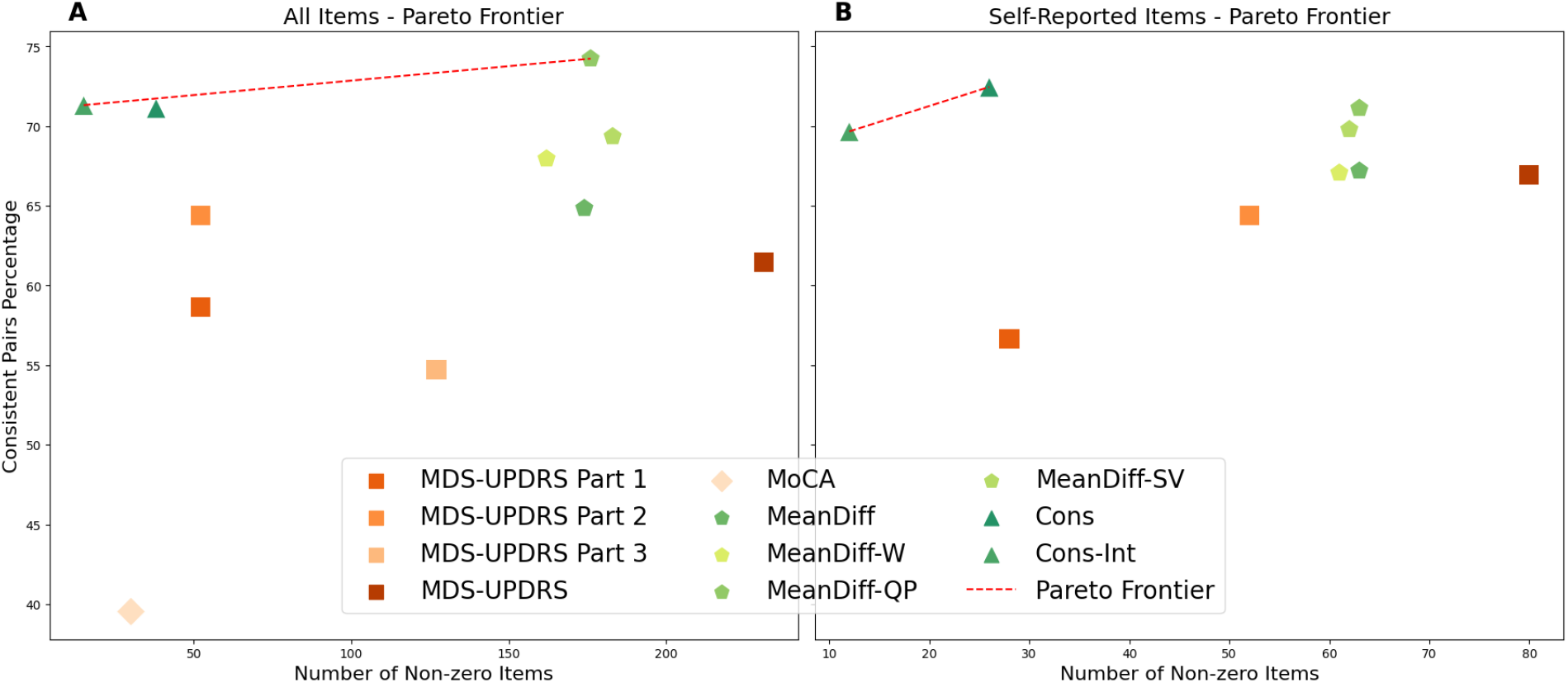
Consistency versus number of items used by each method. Each point represents a method, with the x-axis indicating how many items it includes and the y-axis showing the percentage of visit pairs with increasing scores over time. Methods with the same x-value can be compared by their consistency (higher y is better), while those with the same y-value can be compared by efficiency (lower x is better). An item refers to a single unit increase in a response on the original scale. A. Performance when using all items. B. Performance when using only self-reported items. The red lines indicated the Pareto optimal contour. MDS-UPDRS: Movement Disorder Society’s Unified Parkinson’s Disease Rating Scale. MoCA: Montreal Cognitive Assessment.

In both cases Cons-Int achieved very good consistency, while using a very small number of items. Table 4 shows the learned weights for that solution using only self-reported items. Remarkably, only eleven questions are used, and in ten of those only one threshold value is needed. In the 11th (Getting out of bed) two thresholds are needed. Put differently, this scale uses only twelve self-reported items yet it outperforms the original 200-item MDS-UPDRS. The only scale to achieve higher consistency is Meandiff-QP with 176 items. Supplementary table S3 gives the weights of Const-Int when all items are allowed.

**Table 4.** The scale obtained by Cons-Int using only self-reported items. Only the non zero weights are shown. The final index is obtained by summing the scores for all rows in which the item’s value is equal or larger than the threshold.

| Item | Threshold | Score |
| --- | --- | --- |
| 1.7 Sleep problems | 1 | 13 |
| 1.8 Daytime sleepiness | 1 | 25 |
| 1.10 Urinary problems | 2 | 55 |
| 1.11 Constipation problems | 1 | 43 |
| 1.13 Fatigue | 1 | 30 |
| 2.1 Speech | 1 | 44 |
| 2.2 Saliva and drooling | 1 | 51 |
| 2.9 Turning in bed | 1 | 66 |
| 2.11 Getting out of bed | 1 | 52 |
| 2.11 Getting out of bed | 2 | 45 |
| 2.12 Walking and balance | 2 | 100 |
| 2.13 Freezing | 1 | 67 |

The learned weights for all methods are provided as supplementary files. Supplementary File 1 contains the weights for scales based on all items. Supplementary File 2 contains the weights of scales using only self-reported items.

### 2.3 Additional validation using PPMI

#### 2.3.1 Initiation of symptomatic therapy

Supplementary Table S4 lists the correlation between each tested method and the time to initiation of levodopa. Indeed, we see highly significant negative correlations between the scores and the time difference. Figure 3 shows the results of the best performing method in terms of the significance of correlation in each scenario: MeanDiff-QP using all items and Cons-Int using just self-reported items.

**Figure 3.**
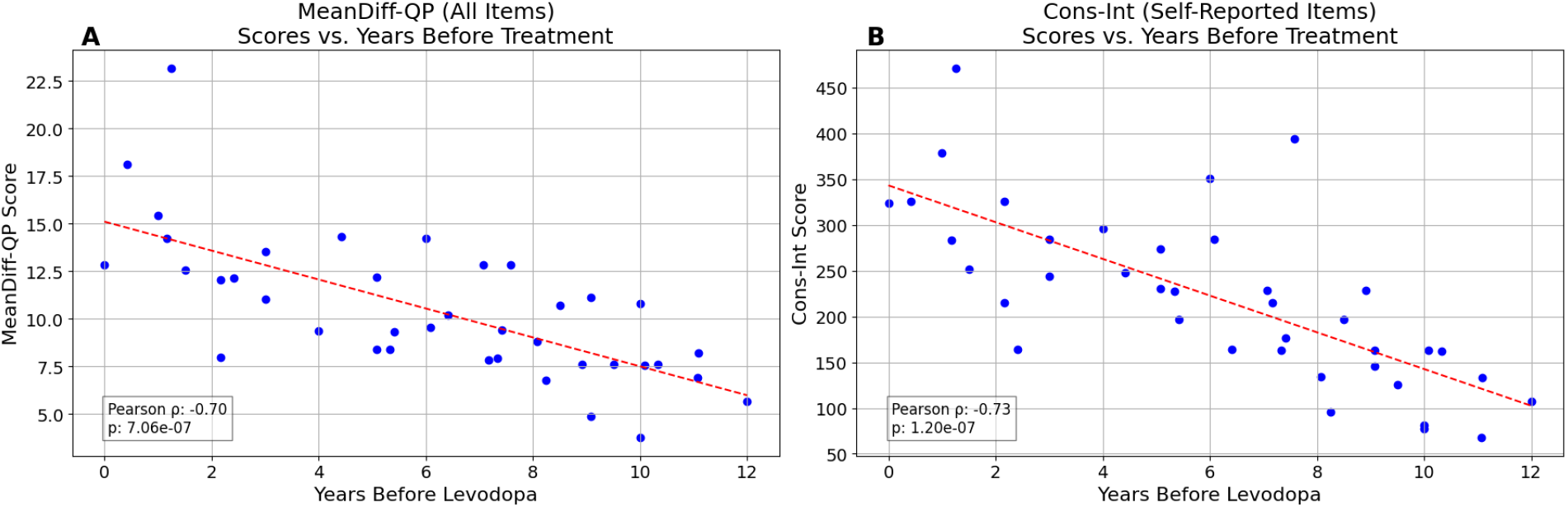
Total scores vs. the number of years prior to initiation of levodopa treatment. A. MeanDiff-QP using all items B. Cons-Int using self-reported items only.

#### 2.3.2 Activities of daily living

When testing the scores correlation with the S&E ADL questionnaire [**13**], the results of all methods were significant (Supplementary Table S5). Figure 4 shows the results for

**Figure 4.**
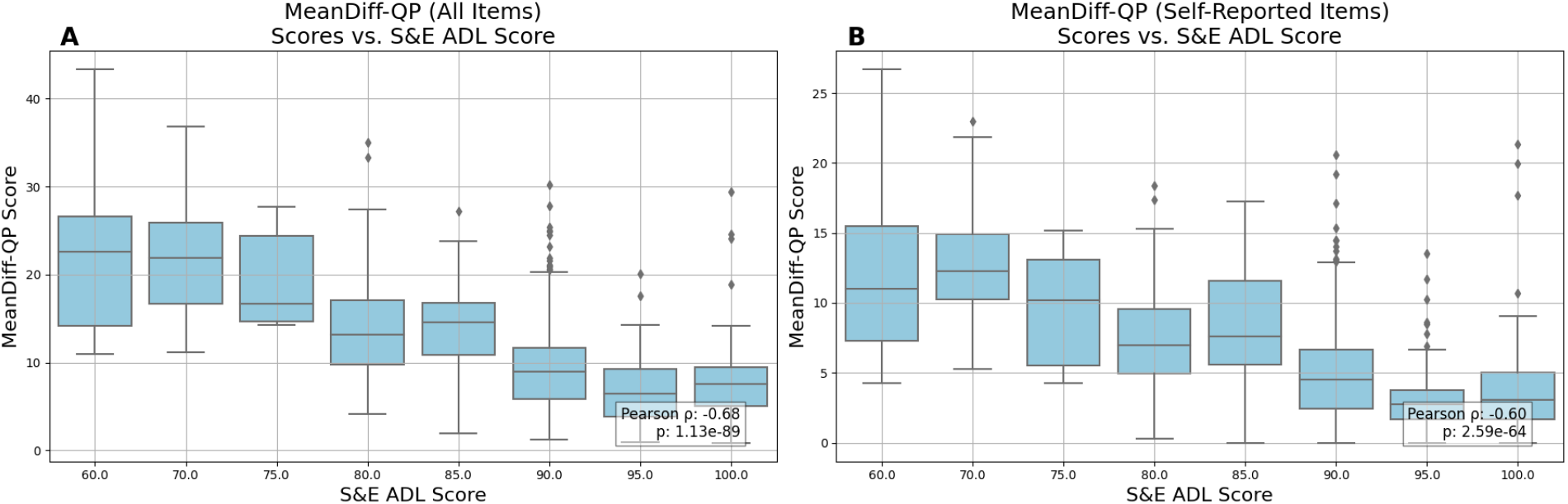
Total scores compared to the S&E ADL scores. A. MeanDiff-QP results when using all items. B. MeanDiff-QP results when using only self-reported items.

MeanDiff-QP, which achieved the highest correlation using all items and the second-best using only self-reported items, surpassed only by MDS-UPDRS Part 2.

Both tests validate the relevancy of our suggested scores, showing high correlations to external data that was not a part of the training process. Many methods outperform the original scales, reaching correlations of −0.73 (p=1.20e-07) with the **Cons-Int** method for time before levodopa treatment and −0.68 (p=1.13e-89) with **MeanDiff-QP** for S&E ADL.

#### 2.3.3 Time to milestone

When measuring the correlation between each index and the time to the first milestone as defined in [**14**], all our scales achieved correlation below −0.41, outperforming the MDSUPDRS. The results for methods using all items can be seen in Figure 5. The correlation coefficients and p values for all methods are available in Supplementary Table S6.

**Figure 5.**
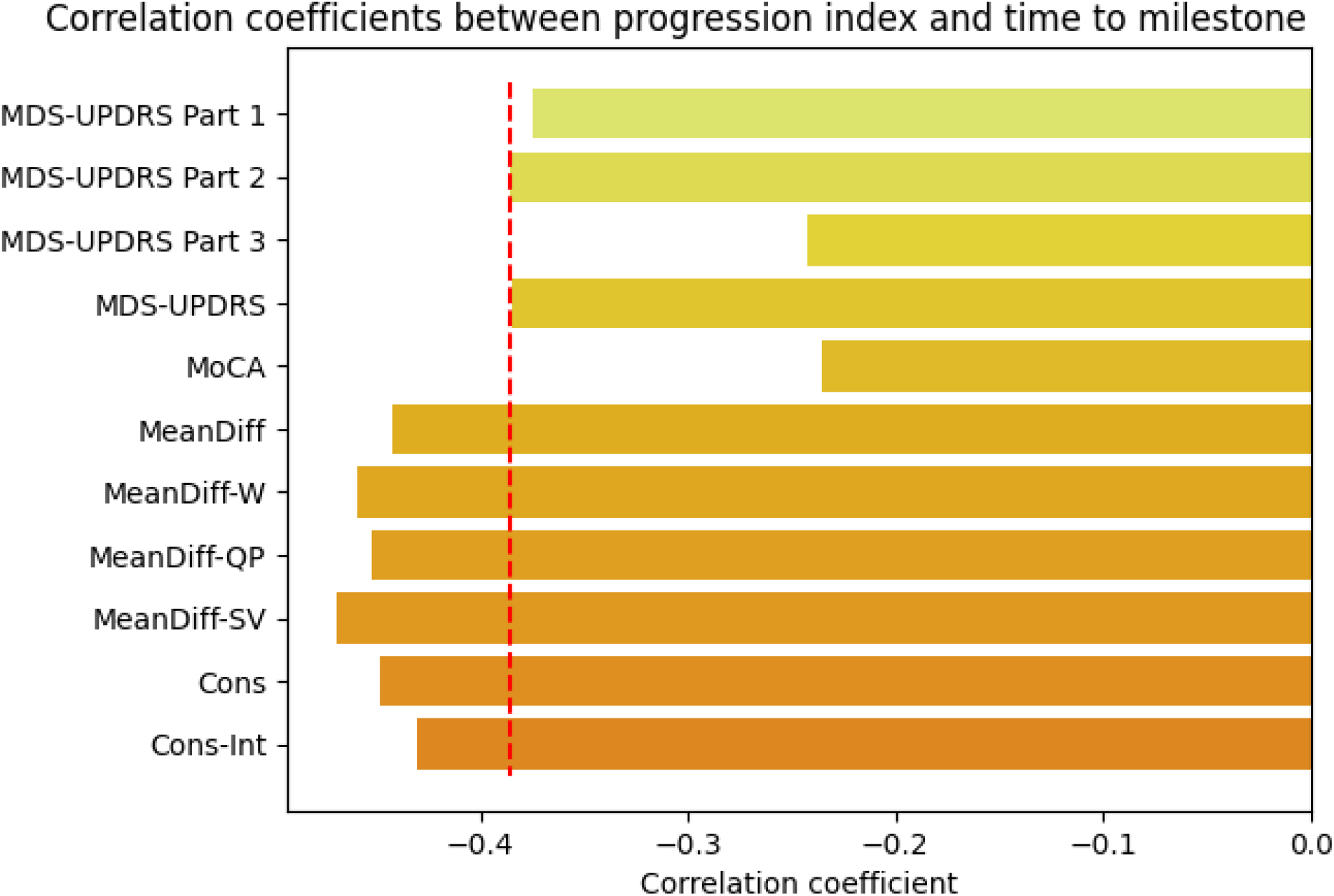
Correlation between the progression index and the time (in month) until the patient reaches at least one of the defined milestones, for each method. All visits after the first visit where a patient reaches a milestone are excluded. The vertical dotted red line indicates the best correlation for a baseline method; our suggested methods exceed this threshold significantly. MDS-UPDRS: Movement Disorder Society’s Unified Parkinson’s Disease Rating Scale. MoCA: Montreal Cognitive Assessment.

### 2.4 External Validation

The participants characteristics of the BeaT-PD cohort after applying the filtering are described in Table 5.

**Table 5.** BeaT-PD participants characteristics. Statistics are shown for the cohort after filtering, consisting of 201 visits of 79 patients. The first five characteristics listed correspond to each patient’s initial visit. MDS-UPDRS: Movement Disorder Society’s Unified Parkinson’s Disease Rating Scale. MoCA: Montreal Cognitive Assessment. H&Y: Hoehn and Yahr

| Characteristic | Mean | Std | Min | Max |
| --- | --- | --- | --- | --- |
| Age (years) | 64.05 | 11.32 | 36 | 86 |
| Disease Duration (years) | 2.82 | 1.94 | 1 | 7 |
| MDS-UPDRS Total Score | 33.18 | 16.69 | 7 | 89 |
| H&Y Stage | 1.68 | 0.57 | 0 | 3 |
| MoCA Total Score | 24.73 | 3.49 | 17 | 30 |
| Gender (M%) | 77.22% |  |  |  |
| Follow-up Time (years) | 3.71 | 1.6 | 1 | 6 |
| Number of Visits | 2.54 | 0.76 | 2 | 5 |

The consistency of all the tested methods on the BeaT-PD cohort is shown in Table 6. Reassuringly, all but one method exceeded the performance of the strongest baseline scale, supporting the robustness of our approach. Validation results using only self-reported items are available in Supplementary B.4.

**Table 6.** The percentage of consistent pairs of visits for each method on the external validation BeaT-PD dataset, evaluated using the scale derived from the PPMI data.

| Method | Consistency (%) |
| --- | --- |
| **MDS-UPDRS P1** | 62.29 |
| **MDS-UPDRS P2** | 65.71 |
| **MDS-UPDRS P3** | 61.71 |
| **MDS-UPDRS** | 65.71 |
| **MoCA** | 54.29 |
| **MeanDiff** | 69.14 |
| **MeanDiff-W** | 70.29 |
| **MeanDiff-QP** | 67.43 |
| **MeanDiff-SV** | **71.43** |
| **Cons** | 64.57 |
| **Cons-Int** | 67.43 |

### 2.5 An online tool

We created an online tool that calculates our progression index using the self-reported answers, available via https://shamir-lab.github.io/MOPS/self_report_short.html. The tool uses the weights of Table 4, normalized so that the range is 0-100.

## 3 Discussion

### Contributions and key findings

We introduced a method for optimizing PD progression indexes by reweighting items and increments in the MDS-UPDRS and MoCA scales. The new indexes have higher precision and efficiency, benefiting both patients and clinicians. Our main findings are: (1) Compared to the current approach of merely summing raw item values, our indexes enhance score consistency with disease progression while maintaining a simple “sum-of-items” format. (2) Indexes based solely on self-reported items perform on par with, or in some cases better than, the full MDS-UPDRS scale, including the parts that are clinician-rated. (3) Indexes using only a few items are almost as good as those based on all items. In particular, eleven self-reported items and twelve weights outperformed the original MDS-UPDRS, which includes 59 items and 236 weights. These findings were corroborated by strong correlations with external progression markers and validated in an external cohort.

### Implications for clinical practice

First, by removing questions that contribute minimally to tracking PD progression, one can focus on the more meaningful indicators of progression without sacrificing diagnostic or prognostic accuracy. Second, the potential to base progression tracking on properly weighted self-reported items alone enables more frequent as well as remote evaluations, offering patients the flexibility to complete assessments at home, while reducing the burden from clinicians. Importantly, our decision to train only on data of patients in ‘ON’ state leads to indexes that are applicable to the real-world daily presentation of patients. Overall, the optimized index could enhance the quality and efficiency of patient care and improve long-term disease management.

### MoCA

The MoCA exhibits a low consistency, and had a minimal contribution to the indexes, likely due to two factors. Many PD patients, particularly in the early stages, do not experience significant cognitive decline. More importantly, MoCA performance is affected by practice effect, where repeated tests lead to improved scores independent of actual cognitive changes [**15**].

### MDS-UPDRS Part 2

It is noteworthy that the score based on part 2 only outperforms the full MDS-UPDRS score. In particular, it outperforms part 3, which is often regarded as the most clinically relevant and reliable. This can be attributed to the influence of medications, which strongly affect the motor symptoms assessed in part 3. Changes in medication or dosage adjustments frequently lead to lower part 3 scores when patients are in the ON state (as in the dataset used here). Additionally, part 2 assessments, being self-reported, avoid the inter-rater variability that affects part 3, reducing measurement noise and improving consistency. Lastly, while part 3 measures the present state, part 2 items usually ask about the last weak, making them less susceptible to symptoms fluctuations.

### Other approaches

One approach relevant to our application is Item Response Theory (IRT) [**16**]. It assumes that each person’s responses are influenced by an underlying trait — in our case, PD severity — and estimates how each item relates to this trait. While IRT has been applied to the MDS-UPDRS [**17**–**22]**, it has some limitations. First, the IRT model assumptions are not fulfilled by the MDS-UPDRS [**23**]. In particular, the assumption that each item is measuring the same trait independently does not hold for the diverse symptoms of PD. Second, IRT does not incentivize sparsity, as it fits the optimal parameters for each item or question separately. Lastly, IRT is primarily designed for cross-sectional data and does not effectively capture changes over time, which are crucial for tracking disease progression.

Additionally, recent studies have proposed re-weighting MDS-UPDRS items using partial least squares regression [**24, 25**]. These methods optimized an internal criterion—the mean-to-standard-deviation ratio—which differs from our focus on consistency. Morinan et al. [**26**] sought to shorten the scale by selecting a subset of eight items suitable for remote monitoring, optimizing explained variance in the process. Unlike our approach, none of these studies allowed for assigning different weights to individual score increments within the same item.

### Early vs. advanced patients

Since PPMI mostly enrolls patients in an early stage of the disease, our data is biased towards early patients; for example, 92.8% of the exams are of patients with H&Y stage ≤ 2. Therefore, the utility of our progression scale will be highest for earlier PD patients, and less informative for more advanced patients. While it is mathematically easy to balance the index and adjust the optimization target to give more weight to more severe patients, we decided against such a change for a few reasons. First, a progression index is much more valuable in earlier stages of the disease, since in later, more severe stages it is easier to identify the progression manifested in a wide range of symptoms. Second, giving more weight to patients with higher H&Y will introduce additional noise and bias, as these stages are characterized by specific aspects of PD, and do not capture the full range of symptoms. Moreover, the H&Y staging itself also exhibits moderate inter-rater reliability [**27**].

### Tremor

Previous research shows that the tremor items in part 3 contain limited information about the underlying state in PD and do not show worsening over time [**28**].Additionally, an IRT scoring of part 3 items gives negative coefficients to the tremor items, claiming they are anti-correlative to the other part 3 items [**29**]. One contributing factor might be that these items are strongly affected by PD medications like levodopa [**30**]. Indeed, in our computational approaches these items usually receive little or no weight, supporting the observation that they are poor indicators of PD progression.

### Computational hardness

Our study explored two ways to rescale the data. The first method focused on an objective that is slightly different from consistency, but it still often led to well-performing scales. We were able to optimize this objective efficiently using fast algorithms. The second method aimed directly at maximizing consistency, but this made the problem much more challenging to solve (in fact, it is proven to be computationally hard—see Supplementary A.5). To tackle it, we used algorithms that can be very slow for large problems. As a result, these algorithms could only find approximately optimal solutions within the available time.

### Limitations and future work

Our study has several limitations. First, we constructed our scales using only data from patients who are drug-naïve or in ON state. This aimed to ensure our results are applicable to patients in their typical daily conditions, where medication are not intentionally withheld. Future studies can use our methodology while focusing on more advanced PD patients who naturally experience frequent OFF-state periods. A more detailed pharmacological profile for each patient—capturing medication types, dosage, and timing —may also allow the model to re-weigh items dominated by temporary symptomatic relief rather than true disease progression.

Second, due to limited computational resources, we split the data into training and test sets but did not allocate a separate validation set for extensive hyperparameter tuning. Instead, for each formulation we tried several parameter values on the training set and took one that performed best. A more systematic approach (e.g., nested cross-validation) using more computation power may lead to better parameter choices and improve the scales.

Finally, including prodromal patients can similarly expand the applicability of our approach, enabling earlier and more nuanced detection of progression trajectories.

### Other domains

While this study focused only on PD, the computational approach and methods provided here can lead to improvement in scales of other diseases or conditions. Examples include Apgar score [**31**] for newborn infants evaluation, the RENAL nephrology scoring system [**32**], the Glasgow Coma Scale [**33**], the Barthel Index for activities of daily living [**34**], the Mini-Mental State Examination for cognition [**35**], the NIH Stroke Scale [**36**] and many others. Such scores are broadly used in healthcare, and improving and simplifying them can increase their utility.

## 4 Methods

### 4.1 Preprocessing

#### 4.1.1 Data

We used data from the Parkinson’s Progression Markers Initiative (PPMI) [**11**] - an international, multi-center longitudinal study aimed at identifying biomarkers of PD progression. In this study, various assessments including MDS-UPDRS are conducted regularly at intervals of 3, 6, or 12 months to track changes in clinical and cognitive status over time (See Supplement A.1 for additional details). While PPMI contains a variety of data types including imaging and genetic data, for constructing the new index we only used MDS-UPDRS [**37**] for summarizing patients’ clinical state and MoCA [**38**] for their cognition.

#### 4.1.2 Filtering

The MDS-UPDRS [**2**] is a 65-item scale divided into four parts (Part I: non-motor experiences of daily living, II: motor experiences of daily living, III: clinician-rated motor examination, IV: motor complications). As our input, we used the 59 questions in parts I, II and III as well as MoCA. While PPMI contains various types of subjects (Healthy, PD, prodromal PD and other disorders) we focused only on PD patients in this analysis, and in particular excluded prodromal patients. We also removed examinations where the rater noted that dyskinesia interfered with the rating.

Since we wanted our tool to be applicable in regular clinical visits, we excluded visits where the PD patients were measured in ‘OFF’ state, as this kind of measurement often requires patients to purposely stop taking their medications and thus introduces undesired burden on them.

Finally, we removed the baseline visit of each patient from our analysis, as we suspect the first visit might be biased due to the Hawthorn effect [**39**], as the act of joining a clinical trial by itself might create some temporal positive “improvement” in the patients state, compared to followup visits.

After filtering the data we had a total of 3,295 examinations for 711 different patients (averaging in 4.63 exams per patient, with median time difference between adjacent visits of 1 year). Note the data is not distributed evenly across PD severity levels, and is heavily biased towards early patients. See supplementary A.1 for the additional details on the PPMI data.

#### 4.1.3 Encoding

To make the data canonic and usable for the next step, we transformed it as follows. First, while MDS-UPDRS gives higher scores for more severe patients, the MoCA score decreases with severity from 30 to 0 - patients get full points for correct answers. To have both monotone increasing with severity, we flipped the values of each MoCA item such that the value is the number of points deducted instead of the number of points gained.

Next, for each question, we assigned a binary variable for each unit increment in the answer. For example, an MDS-UPDRS question that can have an answer between 0 and 4 was transformed into four binary variables *x*_1_, *x*_2_, *x*_3_, *x*_4_, where *x*_*i*_ = 1 if the answer is at least *i*. Hence, the answer 0 is mapped to [0,0,0,0], 1 is mapped to [1,0,0,0], 2 is mapped to [1,1,0,0], 3 is mapped to [1,1,1,0] and 4 is mapped to [1,1,1,1]. This way, for example, the answers to the 59 questions used from the MDS-UPDRS are represented by 236 binary variables. This type of encoding for ordinal data is sometimes referred to as thermometer encoding [**40**] or cumulative binary encoding. By giving non-negative weights to items, *w*_1_, *w*_2_, *w*3, *w*_4_, the score of a question ∑_*i*_ *w*_*i*_ · *x*_*i*_ is monotone non-decreasing: Higher answers are assigned higher scores. The total weighted sum of all answers in a patient’s visit is called its *progression index*.

### 4.2 Evaluation

We split the data into 80% training set and 20% test/evaluation set, such that no patient appeared in both train and test sets. The learning of weights was done only on the training set, and evaluated on the test set.

Our primary metric for assessing the optimized weights was the percentage of visit pairs for the same patient in which the later visit received a higher total score. We call this metric *consistency*. A score with higher consistency is better. We also measured the number of non-zero weights assigned to items. A lower number reflects a simpler scale that is easier to implement.

#### 4.2.1 External validation

We compared the computed progression index against external progression criteria, and tested whether it performs better than the baseline approaches. The first set of criteria were based on data available in PPMI. First, we examined the relationship between a visit’s score and the time elapsed from that visit until the start of levodopa treatment, assuming an effective scale should assign higher scores to patients who are closer to beginning treatment. Second, we checked the scores concordance with the Schwab and England Activities of Daily Living (S&E ADL) scale [**13**], expecting a negative correlation between our disease progression score and the ADL score. Lastly, we used the milestones defined by Brumm et al. [**14**] and checked how well our progression index predicts the time it would take a patient to reach the first milestone. We tested 20 out of the 25 milestones defined in [**14**], for which a sufficiently large fraction of the visits had data. We assumed a good index should exhibit a strong negative correlation, so that higher scores are associated with a shorter time to reaching the first milestone.

Finally, we validated the consistency of our weights against an additional, external cohort of PD patients obtained from the BeaT-PD project (204-16TLV) [**12**]. The BeaT-PD cohort included 300 recently diagnosed patients with PD (mean age at recruitment 61.67±10.34 years with mean disease duration of 2.5±1.1 years) who were clinically and genetically assessed over 5 years. After applying filtering criteria similar to those used for the PPMI dataset, as described in Section 4.1.2 - but without removing baseline visits, to preserve dataset size - we retained 79 patients with a total of 201 visits.

For the validation of the self-report index we applied a milder filtering approach, and did not exclude visits based on MDS-UPDRS part 3 criteria (clinical state or dyskinesia interference), as these are not self-reported measures.

### 4.3 Full index vs self-reported index

We also developed an index that uses only MDS-UPDRS items that are self-reported and do not require a trained rater. This index uses only the items in the patient’s questionnaire (the second half of part I and the entire part II).

### 4.4 Approaches for weights optimization

We developed a variety of formulations for optimizing the weights in the scale. The first set of approaches seek to maximize objective functions that are similar to — but not identical to — the consistency measure, are justified by a solid rationale, and can be optimized efficiently. Empirically, they can be solved to optimality on our data within a few minutes of computation on a standard laptop. These approaches include:

- **MeanDiff** - maximizing the mean difference between pairs of visits of the same patient, across all patients.
- **MeanDiff-W** (Weighted) - similar to the above, but penalizing more for negative differences, corresponding to pairs of visits for which the score decreased. The objective is to maximize the weighted sum of differences.
- **MeanDiff-QP** (Quadratic Penalty) - similar the former but introducing quadratic penalty for decreases - thus penalizing larger decreases more heavily. The objective is to maximize the sum of differences while minimizing the penalty.
- **MeanDiff-SV** (Small Variance) - similar to the MeanDiff approach, with an additional penalty factor measuring the variance of score differences between visits. The objective is to maximize the mean difference while minimizing the differences’ variance, incentivizing stable increases.

For each of the approaches above, we also added an optional regularization term for minimizing the number of non-zero weights, incentivizing sparse solutions. This was both a goal by itself (as discussed earlier), and was also beneficial to prevent overfitting the training data. The full definition for each of these approaches can be found in Supplementary A.3.

Our second set of approaches aim to optimize consistency. They seek weights that will maximize the number of consistent pairs. We considered two variants of this problem: one where weights can have any real value, and one where only integer weights are allowed. We call these formulations **Cons** and **Cons-Int**, respectively. The full definitions are given in Supplementary A.4. The objective functions in these formulations are not convex, so finding the global optimum is computationally harder. We used algorithms that may take exponential time to reach an optimum. In practice, we limited the runtime to a few hours and settled for the best solution found in that time.

### 4.5 Implementation details

All computations were conducted on a system with an AMD EPYC 7702 processor, featuring 128 logical CPUs (64 cores, 2 threads per core) at 2.0 GHz. The machine runs on GNU/Linux 4.15.0-65-generic within an NVIDIA DGX Server environment. Solving Integer Programming and Mixed Integer Programming formulations was done using the Gurobi Optimizer [**41**].

The first set of weights optimization formulations took each up to 30 minutes to complete using just a single thread. The second set of formulations, which aimed to maximize consistency, dealt with hard computational problem and thus was solved using all available cores and were each allotted a 24-hour time limit. Within this timeframe, an optimal solution could not be reached. However, the bound for the gap between the best solution found and the optimal solution ranged between 14.3% and 38.7% across all formulations. These values represent upper bounds, and the actual gaps are likely much smaller.

## 5 Data availability

Access to the PPMI dataset is publicly available upon request at https://www.ppmi-info.org. The BeaT-PD dataset is available from AM upon reasonable request.

## 6 Code Availability

The code developed in this paper is available at https://github.com/Shamir-Lab/MOPS.

## 7 Acknowledgments

This study is dedicated to the late Prof. Nir Giladi, who was a partner and a mentor in this study and sadly passed away during the project’s development.

RS was supported by grants from Israel Science Foundation (grant No. 2206/22) and from the Tel Aviv University Center for AI and Data Science (TAD).

Data used in the preparation of this article was obtained on 2024-08-07 from the Parkinson’s Progression Markers Initiative (PPMI) database (www.ppmi-info.org/access-dataspecimens/download-data), RRID:SCR 006431. For up-to-date information on the study, visit www.ppmi-info.org. The analysis used data openly available from PPMI (Tier 1 Data).

PPMI – a public-private partnership – is funded by the Michael J. Fox Foundation for Parkinson’s Research, and funding partners; including 4D Pharma, Abbvie, AcureX, Allergan, Amathus Therapeutics, Aligning Science Across Parkinson’s, AskBio, Avid

Radiopharmaceuticals, BIAL, BioArctic, Biogen, Biohaven, BioLegend, BlueRock Therapeutics, Bristol-Myers Squibb, Calico Labs, Capsida Biotherapeutics, Celgene, Cerevel Therapeutics, Coave Therapeutics, DaCapo Brainscience, Denali, Edmond J. Safra Foundation, Eli Lilly, Gain Therapeutics, GE HealthCare, Genentech, GSK, Golub Capital, Handl Therapeutics, Insitro, Jazz Pharmaceuticals, Johnson & Johnson Innovative Medicine, Lundbeck, Merck, Meso Scale Discovery, Mission Therapeutics, Neurocrine Biosciences, Neuron23, Neuropore, Pfizer, Piramal, Prevail Therapeutics, Roche, Sanofi, Servier, Sun Pharma Advanced Research Company, Takeda, Teva, UCB, Vanqua Bio, Verily, Voyager Therapeutics, the Weston Family Foundation and Yumanity Therapeutics.

Protocol information for The Parkinson’s Progression Markers Initiative (PPMI) Clinical - Establishing a Deeply Phenotyped PD Cohort AM 3.2. can be found on protocols.io or by following this link: https://dx.doi.org/10.17504/protocols.io.n92ldmw6ol5b/v2. The BeaT-PD project was funded by a grant from Biogen Inc. to Tel Aviv Medical Center in 2016. The sponsor was not involved in the reported analysis, interpretation or presentation of results.

Statistical analysis codes used to perform the analyses in this article are shared on GitHub: https://github.com/Shamir-Lab/MOPS.

## 8 Author contribution

A.B: Conceptualization, data curation, methodology, formal analysis, validation, writing - original draft and edits. R.A.: writing - review. A.M.: Providing validation dataset, writing - review and edits. R.S.: Conceptualization, methodology, formal analysis, writing - review and edits, supervision. The final manuscript was read and reviewed by all authors.

## Appendices

### A Supplementary text

#### A.1 Data

Our data (downloaded from PPMI on August 7 2024) contained information for 1,879 PD patients, 2,089 Prodromal patients and 400 healthy controls. In our analysis we only used the PD patients’ data. Supplementary Table S1 summarizes the data available for these 1,879 PD patients. Part 3 has more visits because patients are often measured twice - in ON and in OFF states. Of the 16,715 visits with Part 3 data, 7,139 were in ON state, 5,158 in OFF and 4,418 were of drug naive patients.

**Table S1.** Statistics on the PD patients in the PPMI. Repeat patients are patients who had more than one visit. MDS-UPDRS: Movement Disorder Society’s Unified Parkinson’s Disease Rating Scale. MoCA: Montreal Cognitive Assessment.

| Exam | Total Visits | Unique Patients | Repeat Patients |
| --- | --- | --- | --- |
| MDS-UPDRS Part 1 | 12,453 | 1,492 | 1,284 |
| MDS-UPDRS Part 2 | 12,467 | 1,496 | 1,284 |
| MDS-UPDRS Part 3 | 16,715 | 1,497 | 1,285 |
| MoCA | 6,544 | 1,709 | 1,096 |

After removing visits in OFF-state and patients with just a single visit, we were left with 4,919 visits of 823 unique patients that contain all MDS-UPDRS parts and MoCA. Removing the baseline and screening visits of all patients left us with 4,269 visits of 763 unique patients. Finally, we filtered visits where dyskinesia interfered with the rating or where critical values were missing or misaligned. The final dataset used in the analysis consisted of 3,295 visits of 711 unique patients (an average of 4.63 visits per patient). Supplementary Table S2 presents the number of visits in each severity level in the final dataset, showing the bias towards early patients.

**Table S2.** Number of visits for each H&Y stage. H&Y: Hoehn and Yahr.

| H&Y Stage | 0 | 1 | 2 | 3 | 4 | 5 |
| --- | --- | --- | --- | --- | --- | --- |
| Number of Visits | 16 | 599 | 2445 | 192 | 39 | 4 |

#### A.2 Mathematical formulations of progression index problem

##### A.2.1 Terminology

We start with some basic definitions needed for formulating our problem.

The questions in the original scale have a range of *k* values of possible answers (For example, 0, …, 4 in the MDS-UPDRS). Without loss of generality we renumber them 1, …, *k*, where higher numbers indicate more severity. Each such question is translated into *k* − 1 binary features called *items*, where item *i* indicates that the answer to the question is at least *i*.

In this convention, a *visit* is a binary vector **v** ∈ {0, 1}^*m*^, where for each item *i, v*_*i*_ = 1 if and only if this item is true for that visit. We denote the *j*-th visit of patient *p* by 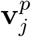. The sequence of visits of patient *p* is denoted by 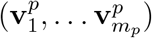, where we assume that for each patient *p m*_*p*_ ≥ 2, and the visits are numbered in increasing time order. A pair of visits 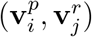 is called *proper* if *p* = *r* and *i < j*. In words, the two visits should be for the same patient and they should be ordered chronologically.

The formulation assigns a *weight w*_*i*_ ∈ ℝ_+_ to each item *i*, together forming a *weight vector* 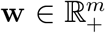. Ensuring that item weights *w*_*i*_ are non-negative and defining the weight of an answer with value *j* as *w*_1_ + … + *w*_*j*_ guarantees that the answer weights are monotone increasing. For a visit **v** and weights **w**, the *score* of the visit is defined as **w** · **v**.

A *longitudinal dataset* is a collection of sequences of visits, one per patient. Formally 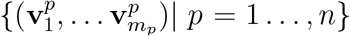. For such dataset, we define *S* as the set of all proper pairs of visits. In other words, 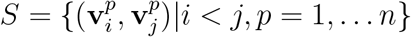.

For a proper pair of visits 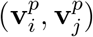 and weights **w**, if 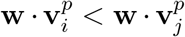 we say that the pair’s order is *consistent with the weights*, or simply that the pair is *consistent*. Note the strict inequality in the last equation. If we allowed instead 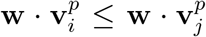, then the weight vector **w** = 0^*m*^ would be a trivial set of weights for which all proper pairs are consistent. We are now ready to define two basic formulations of our problem.

###### Maximum consistency

Given a longitudinal dataset, find a weight vector **w** that maximizes the number of consistent pairs. In other words,

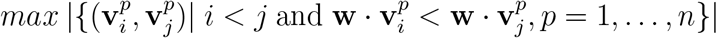

###### Maximum weighted difference

Given a longitudinal dataset, find a weight vector **w** that maximizes the weighted difference across all proper pairs. In other words,

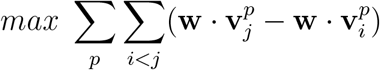

We will introduce several variations of these objectives in the sequel, and also consider a secondary objective of **sparsity**, aiming to reduce the number of items with non-zero weights.

#### A.3 Formulations maximizing the weighted difference

##### A.3.1 Linear Programming

A basic linear programming formulation of the problem is

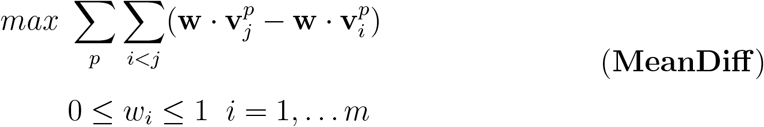

This problem has a closed form solution: The objective is equal to 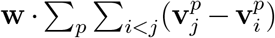. Define 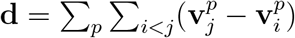. Setting *w*_*i*_ = 1 if *d*_*i*_ *>* 0 and zero otherwise is an optimal solution.

The following formulation takes into account also the solution sparsity:

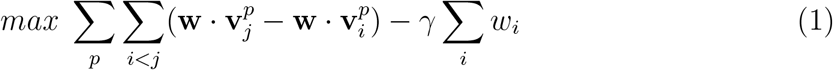

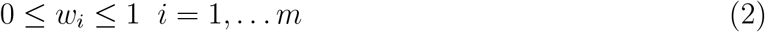

The second term is an L1 regularization of the weights, which incentives sparsity. *γ* is an hyper-parameter that balances between the weighted difference objective and the aim of minimizing the number of used items. This problem too has a closed form solution, since the objective can be written as ∑_*k*_ *w*_*k*_ · *d*_*k*_ − *γ* ∑_*k*_ *w*_*k*_ = ∑_*k*_ *w*_*k*_(*d*_*k*_ − *γ*) so setting *w*_*k*_ = 1 if *d*_*k*_ − *γ >* 0 and zero otherwise is an optimal solution.

###### Variable pair scores

A possible generalization of the first term in the objective is by assigning different values to different pairs: 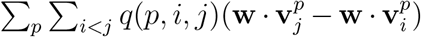. The value *q*(*p, i, j*) of the pair can be used to reduce the weight of visit pairs for patients that have a lot of visits. For example, if patient *p* has *t* visits, then we can make 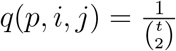 to give each patient equal total weight, or 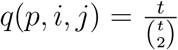 to make the total weight proportional to the number of visits (as opposed to *t*^2^). Alternatively, we can assign different weights to different pairs of visits based on their time span, as larger time gaps are expected to more strongly capture changes in disease severity.

###### Penalizing score drops

This approach is similar to the previous one, but instead of simply maximizing the weighted sum of differences, we would like to punish more heavily inconsistent pairs. We show this for the basic formulation. Denote by *S* the set of all proper visit pairs, where the elements in *S* are the triplets (*p, i, j*) such that *i* and *j* are visits of patient *p* with *j > i*. For each (*p, i, j*) ∈ *S* define nonnegative variables *U*_*p,i,j*_ (for up) and *D*_*p,i,j*_ (down).

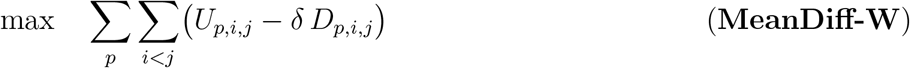

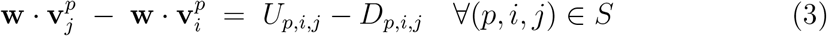

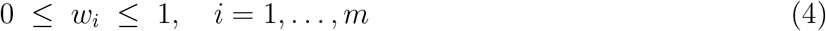

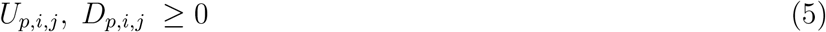

*δ >* 1 is the penalty coefficient for inconsistent pairs.

**Claim:** Any optimal solution of the problem must satisfy:

i. If 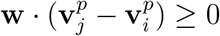, then 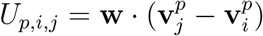 and *D*_*p,i,j*_ = 0.
ii. If 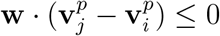, then 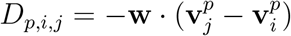 and *U*_*p,i,j*_ = 0.

**Proof:** We prove case (i). The proof of (ii) is analogous. If 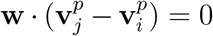 then by (3) *U*_*p,i,j*_ = *D*_*p,i,j*_. The contribution of this triplet (*p, i, j*) to the objective is *D*_*p,i,j*_ − *δD*_*p,i,j*_, which is negative since *δ >* 1 unless *U*_*p,i,j*_ = *D*_*p,i,j*_ = 0.

If 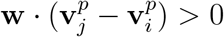, suppose 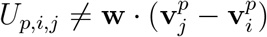. Define 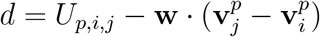. To satisfy (3), we get *D*_*p,i,j*_ = *d. d* ≥ 0 due to the non-negativity constraints. Assume by contradiction that *d >* 0. The objective function then changes by:

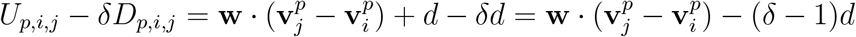

Since *δ >* 1 and *d >* 0, we get a strictly worse objective value than if *d* = *D*_*p,i,j*_ = 0, in contradiction to the assignment being optimal. ▪

##### A.3.2 Quadratic Programming

Similarly to the linear programming approach, we can also introduce quadratic terms in the objective - thus formulating a quadratic programming problem.

###### Squaring the changes

This approach simply squares *U*_*p,i,j*_ and *D*_*p,i,j*_, thus giving more weight to the big changes compared to the smaller ones:

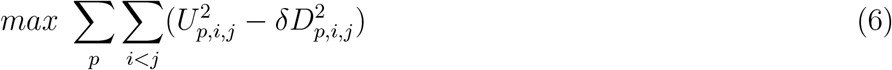

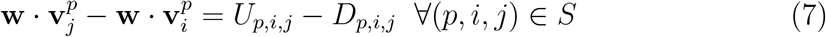

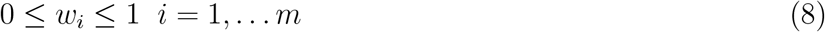

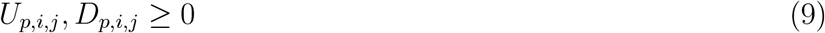

###### Penalizing drops quadratically

Instead of squaring both terms, here we do so just for the drops - so the loss from a drop is bigger than the gain from an increase of the same size. This steers the solution toward greater consistency. We do it by replacing the objective with:

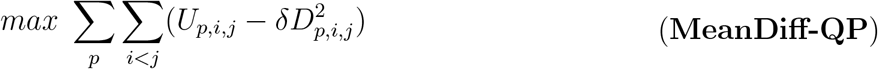

###### Mixing linear and quadratic penalties

The caveat of the last approach is that it is tolerant to small decreases. To avoid that, we mix both linear penalty and quadratic penalties for drops, using two coefficients:

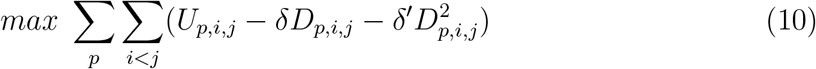

In our tests we used version (**MeanDiff-QP**), as we preferred a minimal amount of hyper-parameters.

###### Reducing variance

Another approach utilizing quadratic programming adds a penalty for the variance of score differences. Denote by 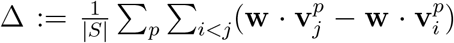 the mean difference in the total score between pairs of visits. The new objective function is:

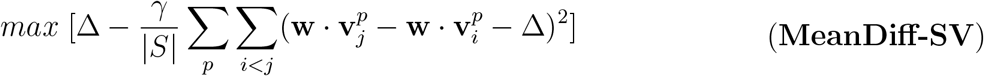

Where again *γ* is an hyper-parameter that balances between the weighted difference objective and the objective of the differences being more stable.

#### A.4 Formulations maximizing consistency

In this section we describe formulations that aim to find integer weights that directly maximize the consistency.

##### Matrix representation

Recall that *S* is the set of all proper visit pairs, where the elements in *S* are the triplets (*p, i, j*) such that *i* and *j* are visits of patient *p* with *j > i*. Denote *s* := |*S*|. We define a matrix of differences **A** ∈ {−1, 0, 1}^*s×m*^, such that for every triplet *S*_*l*_ = (*p, i, j*) and item *a* ∈ {1, …, *m*}, we have 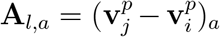. In words, **A**_*l,a*_ is the difference in the value of item *a* between visits *j* and *i* of patient *p*. Since items are binary **A**’s entries are 1, 0 or −1.

##### Integer Programming (IP)

We define two boolean vectors of indicators **I**^+^, **I**^−^ ∈ {0, 1}^*s*^, and use the following IP formulation:

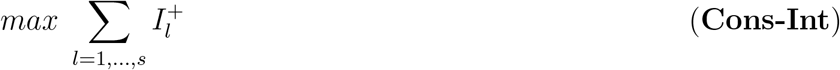

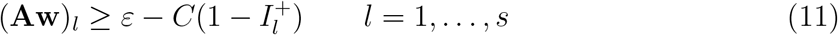

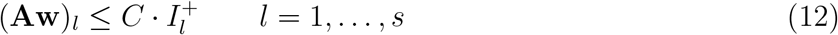

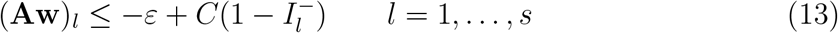

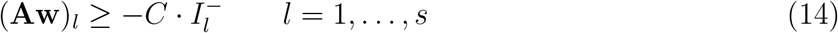

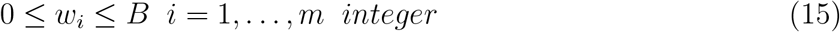

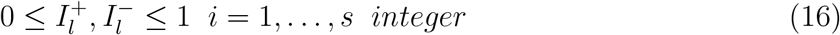

Where 0 *< ϵ* ≪ 1 is a sufficiently small constant, *B* is an upper bound on weight values, and *C* is a large constant such that *C > B*· *m*+*ε*. We call this problem, which maximizes consistency and requires weight integrality **Cons-Int**. The version where (15) is changed so that weights *w*_*i*_ can be real valued is called **Cons**.

**Lemma:** Let **w, I**^+^, **I**^−^ be a feasible solution of the problem. Then for every *l* = 1, …, *s*:

1. (**Aw**)_*l*_ *>* 0 if and only if 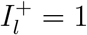.
2. (**Aw**)_*l*_ *<* 0 if and only if 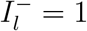.
3. (**Aw**)_*l*_ = 0 if and only if 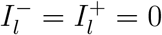.

**Proof:** (1) Assume 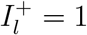. From (11) we get (**Aw**)_*l*_ ≥ *ε*−*C*(1 − 1) = *ε*. Since 0 *< ϵ <* 1, (**Aw**)_*l*_ *>* 0. In the other direction, assume (**Aw**)_*l*_ *>* 0 and 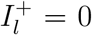. Then from (12) we get 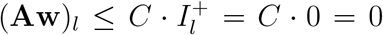, a contradiction. (2) Assume 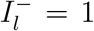. From (13) we get (**Aw**)_*l*_ ≤ −*ε* + *C*(1 − 1) = −*ε*. Since 0 *< ϵ <* 1, (**Aw**)_*l*_ *<* 0. In the other direction, assume (**Aw**)_*l*_ *<* 0 and 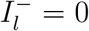. Then from (14) we get 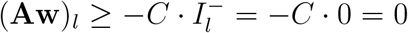, a contradiction. (3) follows from (1) and (2). ▪

**Claim:** The solution to Cons-Int maximizes consistency.

**Proof:** By the lemma, our objective is equivalent to consistency. Denote by **ŵ** an integer vector of weights 0 ≤ *ŵ*_*k*_ ≤ *B* that achieves optimal consistency. We will show that it corresponds to a feasible solution.

For every consistent pair *S*_*l*_ according to **ŵ** we assign 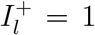 and 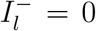. For every inconsistent pair *S*_*l*_ we assign 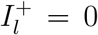, and assign 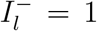 if (**Aŵ**)_*l*_ *<* 0 or 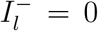 if (**Aŵ**)_*l*_ = 0. We claim (**ŵ**, Î^+^, Î^−^) is a feasible solution. Constraints (15) are satisfied by assumption, and (16) by construction. Assume first *S*_*l*_ is consistent, i.e. 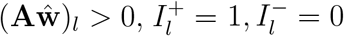. The constraint (11) holds since **ŵ** is an integer vector and **A** ∈ {−1, 0, 1}^*s×m*^, so (**Aŵ**)_*l*_ ≥ 1 *> ε*. Constraint (12) holds since (**Aŵ**)_*l*_ ≤ *mB < C*. Constraint (13) holds since (**Aŵ**)_*l*_ ≤ *mB < C* − *ε*. Constraint (14) holds since (**Aŵ**)_*l*_ *>* 0.

Assume now *S*_*l*_ is inconsistent and decreasing, i.e. 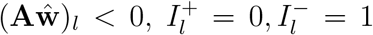. The constraint (11) holds since (**Aŵ**)_*l*_ ≥ −*mB >* −*C* + *ε*. (12) holds since (**Aŵ**)_*l*_ ≤ 0. (13) holds since (**Aŵ**)_*l*_ *<* 0 ((**Aŵ**)_*l*_ ≤ −1). (14) holds since (**Aŵ**)_*l*_ ≥ −*mB >* −*C* + *ε >* −*C*.

Finally, assume *S*_*l*_ is inconsistent and unchanged, i.e. 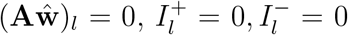. The constraint (11) holds since (**Aŵ**)_*l*_ ≥ −*mB >* −*C* + *ε*. (12) holds since (**Aŵ**)_*l*_ ≤ 0. (13) holds since (**Aŵ**)_*l*_ ≤ *mB < C* − *ε*. (14) holds since (**Aŵ**)_*l*_ ≥ 0. ▪

##### Sparsity

To encourage a sparse solution, we can introduce a regularization term to the objective, as before:

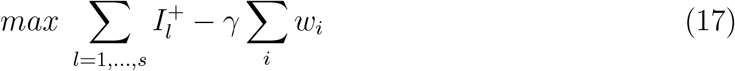

This drives down the sum of the weights, but not sparsity per se. The following IP formulation achieves this goal. To penalize the number of non-zero weights, we introduce helper boolean variables **z** ∈ {0, 1}^*m*^ and formulate the problem as

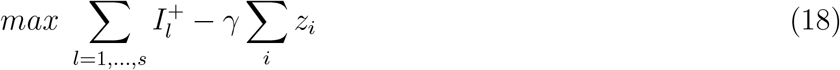

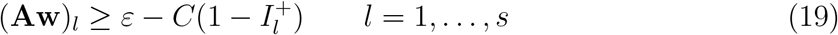

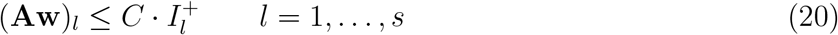

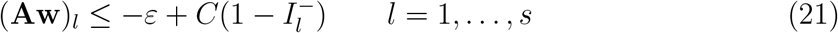

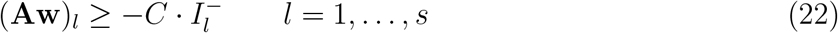

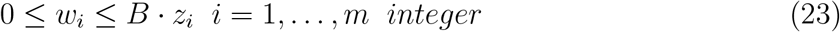

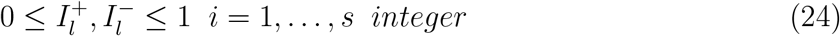

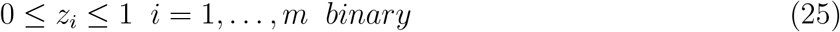

Where again *γ >* 0 balances between consistency and sparsity.

**Claim:** An optimal solution of the system must satisfy ∑ _*i*_ **z**_*i*_ = |{*i*|**w**_*i*_ *>* 0}|

**Proof:** We claim *z*_*i*_ = 1 if and only if *w*_*i*_ *>* 0, from which the claim follows. If *w*_*i*_ *>* 0 then it must be that *z*_*i*_ = 1, to satisfy 23. If *z*_*i*_ = 1 but *w*_*i*_ = 0, then assigning *z*_*i*_ = 0 will increase the objective value by *γ* without invalidating any constraint, in contradiction to the solution’s optimality. ▪

Thus, the penalty term is exactly *γ* times the number of non-zero weights.

##### Mixed Integer Programming

We can relax the integrality constraints for the weights *w*_*i*_, and allow them to have real values instead. This yields a Mixed Integer Programming (MIP) formulation, which can be solved similarly to the previous one. In our tests we tried both approaches, and included a regularization term for sparsity.

##### An alternative objective

By definition, when optimizing for consistency we do not differentiate between the cases where 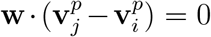 and 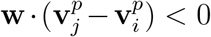. However, no change is preferred over negative change when pursuing a monotonic score. This can be achieved using a similar formulation, by adding a penalty term for the number of strictly negative decreases, with a balance parameter *γ*:

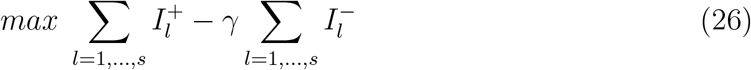

We note that a sparsity term can also be added to the objective.

#### A.5 Hardness of the computational problem

In this section we show that the maximum consistency problem is NP-hard.

First, recall the Partial Maximum Feasible Subsystem problem (Partial Max-FS, also called Constrained Max-FS) [**42, 43**]. In Partial Max-FS we are given a set of linear inequality constraints, where some of them are called hard constraints and the rest are called soft constraints. We wish to find a largest cardinality subset of the constraints containing all the hard constraints and some of the soft constraints that is feasible. Partial Max-FS is NP-Hard and also hard to approximate efficiently [**44**]. It is also NP-hard if the variables are integer or binary, and if the inequalities are strict (*<*) [**43**].

Observe that the consistency maximization problem is a special case of Partial Max-FS. In our case, the variables are binary, the non-negativity constraints are hard, and set of soft constraints is the set of examination pairs, where for each pair we want the score of the later examination to be higher. The optimal weights are those that maximize the number of constraints that are satisfied.

**Theorem:** The consistency maximization problem is NP-Hard, even when *B* = 1 (i.e., **w** ∈ {0, 1}^*m*^).

**Proof:** Observe first that any matrix **A** ∈ {−1, 0, 1}^*s×m*^ can be viewed as a matrix of differences of a set of visits as defined in Section A.4. This follows by forming a dataset with *s* patients where each patient *i* has exactly two visits, and the coordinate-wise differences between the item values in the two visits match the values in row **A**_*i*,·_. By the observation, we can conveniently discuss the problem in terms of constraints, where the weights are the variables.

We show a reduction from the 3-SAT problem. Given a 3-SAT instance with *k* variables and *n* clauses, we construct an instance of the consistency maximization problem as follows:

##### Variable Constraints

Set *m* = 2·*k*, where for each variable *x*_*i*_, the index 2*i*−1 corresponds to the positive literal *x*_*i*_, and the index 2*i* corresponds to the negation ¬*x*_*i*_. For each variable *x*_*i*_, introduce two inequalities:

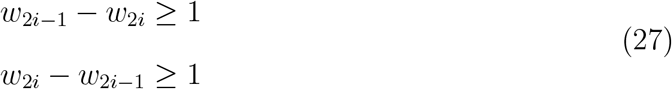

#### Observation

If *w*_2*i*−1_ = 1 and *w*_2*i*_ = 0, or *w*_2*i*−1_ = 0 and *w*_2*i*_ = 1, then exactly one of the inequalities (27) holds. If *w*_2*i*−1_ = *w*_2*i*_ = 1, or *w*_2*i*−1_ = *w*_2*i*_ = 0, then none holds.

##### Clause Constraints

For each clause *C*_*l*_ in the 3-SAT formula, introduce an inequality corresponding to the literals in the clause. Specifically:

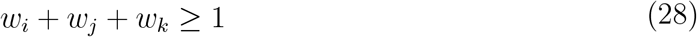

where *i, j, k* are the indices corresponding to the literals in the clause.

Clearly, this reduction is polynomial in the size of the 3-SAT input. Note that since the variables *w*_*i*_ ∈ {0, 1} and all the coefficients in (27) and (28) are 0,1, or −1, the constraints ≥ 1 are equivalent to the ≥ *ϵ* constraints that we had in the Cons-Int formulation.

We claim that the 3-SAT instance is satisfiable if and only if there exists **w** ∈ {0, 1}^*m*^ that achieves consistency in exactly *n* + *k* vectors of differences.

For proof, by the observation, out of each variable-related pair (27) at most one constraint can be satisfied by any assignment. Therefore, the maximal possible number of simultaneously feasible inequalities is *n* + *k*.

Assume the 3-SAT instance is satisfiable. Then there exists an assignment of the *k* variables that satisfies all *n* clauses. Construct the weight vector **w** as follows: For each variable *x*_*i*_: If *x*_*i*_ is True, set *w*_2*i*−1_ = 1 and *w*_2*i*_ = 0. If *x*_*i*_ is False, set *w*_2*i*−1_ = 0 and *w*_2*i*_ = 1. By the observation, exactly *k* inequalities corresponding to the variables are consistent. Since all clauses are satisfied by the assignment, each clause inequality has at least one corresponding index in **w** set to 1, making all *n* clause inequalities consistent. Therefore, the total number of satisfied constraints is *n* + *k*.

Conversely, assume there exists a weight vector **w** such that *n* + *k* inequalities hold. Since there are *k* variable-related pairs, at least *k* of the satisfied inequalities correspond to the variable constraints. By the observation, for each variable *x*_*i*_, exactly one of the inequalities (27) holds. If *w*_2*i*−1_ = 1 and *w*_2*i*_ = 0, set *x*_*i*_ to be True. If *w*_2*i*−1_ = 0 and *w*_2*i*_ = 1, set *x*_*i*_ to be False. Since the total number of satisfied inequalities is *n* + *k*, it follows that all *n* clause inequalities hold as well. By construction, a clause vector is consistent if and only if at least one of its corresponding literals is True. Therefore, the assignment satisfies all clauses. ▪

## APPENDIX B

### B Figures and tables

#### B.1 The weights learned by each approach

Supplementary File 1 contains the weights for scales based on all items. Supplementary File 2 contains the weights of scales using only self-reported items. For each method, the values were normalized to sum to 100. Note that for MoCA items the values were flipped, so for example a threshold of 1 means 1 point below the maximal possible score.

**Table S3.** The scale obtained by Cons-Int when all items can be used. Only non zero weights are shown. The index is obtained by summing the scores for all rows where the item’s value is equal or larger than the threshold. MoCA: Montreal Cognitive Assessment.

| Item | Threshold | Score |
| --- | --- | --- |
| 1.1 Cognitive impairment | 1 | 16 |
| 1.7 Sleep problems | 1 | 9 |
| 1.7 Sleep problems | 2 | 25 |
| 2.2 Saliva and drooling | 2 | 14 |
| 2.3 Chewing and swallowing | 1 | 10 |
| 2.3 Eating tasks | 1 | 13 |
| 2.8 Doing hobbies and other activities | 1 | 7 |
| 2.9 Turning in bed | 1 | 27 |
| 2.12 Walking and balance | 2 | 47 |
| 2.13 Freezing | 1 | 35 |
| 3.4b Finger tapping - Left hand | 1 | 13 |
| 3.7b Toe tapping - Left hand | 1 | 7 |
| 3.13 Posture | 1 | 19 |
| 3.13 Posture | 3 | 100 |
| MoCA - Clock hands | <1 (fail) | 15 |

#### B.2 Comparison of the indexes to external scales

**Table S4.** Correlation between the score of each method and the time to Levodopa. Results are shown for scores that use all items and for scores that use self-reported items only. p-values are calculated using Pearson’s *ρ*. MDS-UPDRS: Movement Disorder Society’s Unified Parkinson’s Disease Rating Scale. MoCA: Montreal Cognitive Assessment.

| Method | All Items | Self-Reported Items |
| --- | --- | --- |
| MDS-UPDRS Part 1 | -0.31 (p=5.09e-02) | -0.25 (p=1.19e-01) |
| MDS-UPDRS Part 2 | -0.67 (p=3.83e-06) | -0.67 (p=3.83e-06) |
| MDS-UPDRS Part 3 | -0.46 (p=3.44e-03) | NA |
| MDS-UPDRS | -0.63 (p=2.07e-05) | -0.64 (p=1.15e-05) |
| MoCA | 0.41 (p=1.04e-02) | NA |
| MeanDiff | -0.62 (p=2.62e-05) | -0.60 (p=4.57e-05) |
| MeanDiff-W | -0.64 (p=9.98e-06) | -0.61 (p=3.19e-05) |
| MeanDiff-QP | **-0.70** (p=7.06e-07) | -0.67 (p=3.30e-06) |
| MeanDiff-SV | -0.69 (p=1.51e-06) | -0.61 (p=4.33e-05) |
| Cons | -0.62 (p=2.73e-05) | -0.68 (p=1.74e-06) |
| Cons-Int | -0.60 (p=6.02e-05) | **-0.73** (p=1.20e-07) |

#### B.3 Correlation of the indexes with the time to first milestone

**Table S5.** Correlation between the score of each method and S&E ADL. Results are shown for scores that use all items and for scores that use self-reported items only. p-values are calculated using Pearson’s *ρ*. MDS-UPDRS: Movement Disorder Society’s Unified Parkinson’s Disease Rating Scale. MoCA: Montreal Cognitive Assessment.

| Method | All Items | Self-Reported Items |
| --- | --- | --- |
| MDS-UPDRS Part 1 | -0.42 (p=3.32e-29) | -0.31 (p=6.62e-16) |
| MDS-UPDRS Part 2 | -0.62 (p=3.51e-69) | **-0.62** (p=3.51e-69) |
| MDS-UPDRS Part 3 | -0.40 (p=2.43e-26) | NA |
| MDS-UPDRS | -0.59 (p=1.33e-62) | -0.57 (p=2.80e-57) |
| MoCA | -0.41 (p=2.90e-27) | NA |
| MeanDiff | -0.65 (p=3.64e-78) | -0.56 (p=8.12e-54) |
| MeanDiff-W | -0.65 (p=2.55e-78) | -0.56 (p=8.53e-55) |
| MeanDiff-QP | **-0.68** (p=1.13e-89) | -0.60 (p=2.59e-64) |
| MeanDiff-SV | -0.62 (p=4.14e-69) | -0.54 (p=1.31e-50) |
| Cons | -0.64 (p=7.17e-77) | -0.58 (p=1.14e-60) |
| Cons-Int | -0.57 (p=8.55e-58) | -0.52 (p=2.20e-45) |

#### B.4 External validation using self-reported items

Table S7 shows the consistency results on the external cohort when using only the self-reported items. In this analysis we did not filter by clinical state or presence of dyskinesia, which are relevant to part 3 of MDS-UPDRS. Only one of our suggested methods fell behind the best baseline method. Also, the best performing method using all items, MeanDiff-SV, is ranked second-best in the self-report-only setting.

**Table S6.** Correlation between each method’s score and the time to first milestone. Results are shown for scores that use all items and for scores that use self-reported items only. p-values are calculated using Pearson’s *ρ*. MDS-UPDRS: Movement Disorder Society’s Unified Parkinson’s Disease Rating Scale. MoCA: Montreal Cognitive Assessment.

| Method | All Items | Self-Reported Items |
| --- | --- | --- |
| MDS-UPDRS Part 1 | -0.37 (p=3.75e-56) | -0.33 (p=4.54e-44) |
| MDS-UPDRS Part 2 | -0.39 (p=9.49e-60) | -0.39 (p=9.49e-60) |
| MDS-UPDRS Part 3 | -0.24 (p=1.39e-23) | NA |
| MDS-UPDRS | -0.38 (p=2.00e-59) | -0.41 (p=9.70e-68) |
| MoCA | -0.24 (p=2.98e-22) | NA |
| MeanDiff | -0.44 (p=4.69e-80) | **-0.42** (p=3.84e-71) |
| MeanDiff-W | -0.46 (p=7.03e-87) | **-0.42** (p=5.53e-72) |
| MeanDiff-QP | -0.45 (p=6.44e-84) | **-0.42** (p=1.94e-71) |
| MeanDiff-SV | **-0.47** (p=2.97e-91) | **-0.42** (p=2.87e-71) |
| Cons | -0.45 (p=1.54e-82) | **-0.42** (p=5.42e-73) |
| Cons-Int | -0.43 (p=1.80e-75) | -0.41 (p=4.55e-67) |

**Table S7.** Percentage of consistent visit pairs for each method on the external validation dataset, evaluated with PPMI-derived weights based solely on self-reported items. MDS-UPDRS: Movement Disorder Society’s Unified Parkinson’s Disease Rating Scale. MoCA: Montreal Cognitive Assessment.

| Method | Consistency (%) |
| --- | --- |
| **MDS-UPDRS P1** | 51.57 |
| **MDS-UPDRS P2** | 68.85 |
| **MDS-UPDRS** | 68.06 |
| **MeanDiff** | 69.63 |
| **MeanDiff-W** | 68.85 |
| **MeanDiff-QP** | 70.94 |
| **MeanDiff-SV** | 71.99 |
| **Cons** | **74.61** |
| **Cons-Int** | 66.23 |

